# Willingness to receive vaccination against COVID-19: results from a large nationally representative Australian population survey

**DOI:** 10.1101/2021.07.22.21260551

**Authors:** Sharon R Davis, Rosario D Ampon, Leanne M Poulos, Guy B Marks, Brett G Toelle, Helen K Reddel

## Abstract

We surveyed 10,024 Australians regarding COVID-19 vaccine willingness. Overall, 59.9% indicated yes, 13.9% no and 26.3% unsure/don’t know. Vaccine willingness was higher in males, and increased with increasing education and socioeconomic advantage. Results contrast with earlier, smaller Australian surveys regarding vaccination willingness and confirm the need for targeted vaccination information.

---

The success of COVID-19 vaccination programmes depends on vaccination acceptance, which varies by local context^1^ and over time. We surveyed a nationally representative population of 10,024 Australian adults between 8/12/2020 and 7/01/2021, when Australia had few active COVID-19 cases and vaccination rollout was imminent. We asked, “*If a COVID-19 vaccine were to become available, would you get vaccinated?*” Overall, 59.9% of respondents indicated “yes”, 13.9% “no” and 26.3% “unsure/don’t know”. Females were less likely than males (54.2% vs 66.0%, age-adjusted odds ratio (OR [95% confidence limit]) 0.60 [0.52-0.65]) to say “yes”, as were people speaking a language other than English at home (57.0% vs 60.5%, age-adjusted OR 1.0 [0.9-1.12]. Vaccination willingness increased with years of education and socioeconomic advantage (Table 1) and was much higher among those who had an influenza vaccination in the previous year than those who had not (72.4% vs 41.7%; age-adjusted OR 3.44 [3.15-3.76]).

**Table 1.**
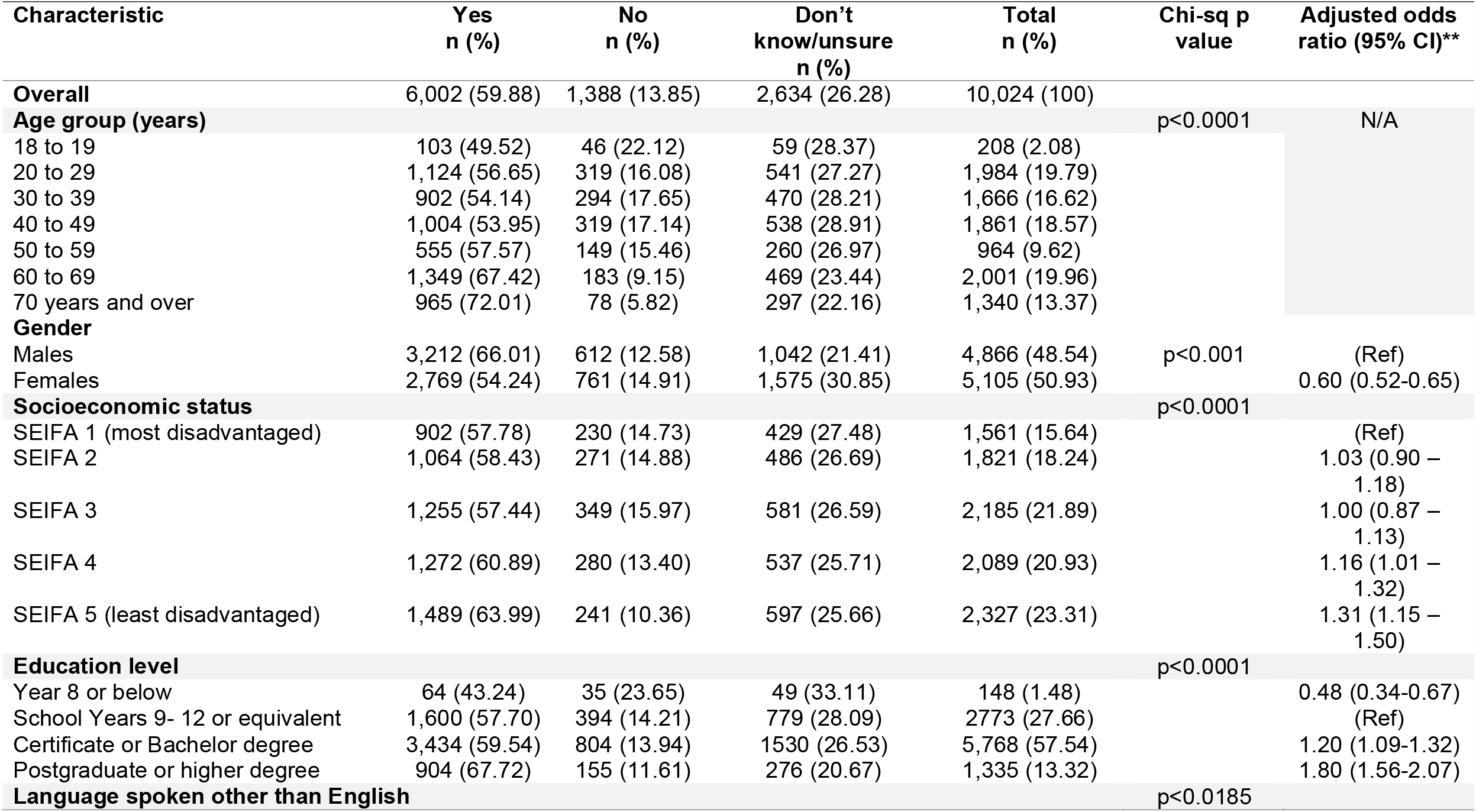
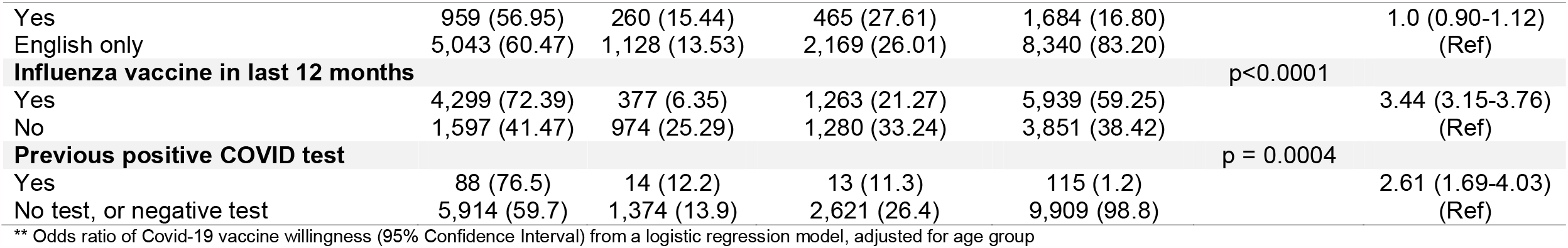
Vaccination willingness

Of those reporting a previous positive COVID-19 test (4.3% of n=2677 tested), 76.5% indicated they would get vaccinated, 12.2% would not and 11.3% were not sure (age-adjusted OR 2.61 [1.69-4.03]).

Qualitative analyses of free text explanations showed that vaccination willingness reflected the desire to be protected at a personal or community level; comments about reducing communal risk included “the right thing to do” “and “herd immunity”. Other reasons were to enable travel, visit or care for others, and virus elimination. Vaccine hesitancy or apathy responses included that vaccine development was “too rushed”, more testing for efficacy and/or safety was needed; healthcare professional advice about vaccine effects on other conditions or medications was needed; and not being in a perceived susceptible group or location.

These results contrast with earlier, smaller Australian surveys^2,3^. Vaccination willingness was reported as 85.8% in April 2020^2^ (n=4362, with vaccination reluctance associated with low health literacy and lower education level) and 75% in June 2020 (n=2018)^3^. These suggest a progressive decline in vaccination willingness over time; Biddle et al^4^ reported that at an individual level, 31.9% of the 2737 Australians they surveyed became less willing to get the vaccine between August 2020 and January 2021.Vaccine hesitancy may have increased since then with recent publicity about adverse effects such as clotting.

Our findings confirm the need for targeted COVID-19 vaccination information for groups including women, and people with lower education levels, but also the need to identify attitudes, beliefs and perceptions to be addressed to achieve widespread community acceptance of COVID-19 vaccination in areas or populations, or at times during which there may be lower perceived risk of COVID-19 infection.

## Data Availability

Data can be made available after Human Ethics Review Committee approval of an analyses plan.

